# Quantifying the risks versus benefits of the Pfizer COVID-19 vaccine in Australia: a Bayesian network analysis

**DOI:** 10.1101/2022.02.07.22270637

**Authors:** Jane E Sinclair, Helen J Mayfield, Kirsty R Short, Samuel J Brown, Rajesh Puranik, Kerrie Mengersen, John CB Litt, Colleen L Lau

## Abstract

The Pfizer COVID-19 vaccine is associated with increased myocarditis incidence. Constantly evolving evidence regarding incidence and case fatality of COVID-19 and myocarditis related to infection or vaccination, creates challenge for risk-benefit analysis of vaccination programs. Challenges are complicated further by emerging evidence of waning vaccine effectiveness, and variable effectiveness against variants. Here, we build on previous work on the COVID-19 Risk Calculator (CoRiCal) by integrating Australian and international data to inform a Bayesian network that calculates probabilities of outcomes for the Delta variant under different scenarios of Pfizer COVID-19 vaccine coverage, age groups (≥12 years), sex, community transmission intensity and vaccine effectiveness. The model estimates that in a population where 5% were unvaccinated, 5% had one dose, 60% had two doses and 30% had three doses, the probabilities of developing and dying from COVID-19-related myocarditis were 239-5847 and 1430-384,684 times higher (depending on age and sex), respectively, than developing vaccine-associated myocarditis. For one million people with this vaccine coverage, where transmission intensity was equivalent to 10% chance of infection over two months, 68,813 symptomatic COVID-19 cases and 981 deaths would be prevented, with 42 and 16 expected cases of vaccine-associated myocarditis in males and females, respectively. The model may be updated to include emerging best evidence, data pertinent to different countries or vaccines, and other outcomes such as long COVID.

## 1. INTRODUCTION

In December 2020, the Pfizer vaccine (BNT162b2; Cormirnaty) became the first COVID-19 vaccine to be authorized for public use [1], and has since had more than 1.5 billion doses delivered to 131 countries [2,3]. In June 2021, reports linking the Pfizer vaccine to myocarditis, especially in male adolescents and young adults, started to emerge in Israel [4] and the USA [5]. Despite low case numbers, this association informed government policies surrounding a slower vaccine rollout in younger age groups around the world [6]. Furthermore, intense media focus on this rare adverse event may have contributed to an increase in vaccine hesitancy in younger age groups [7], especially in Australia where it was the only COVID-19 vaccine recommended for those aged under 60 years at the time [8].

Having access to transparent information on the risks and benefits based on the current best available evidence is crucial for individuals to make an informed decision on whether or not to get vaccinated [9,10], and also for informing public health policy. The Australian Technical Advisory Group on Immunisation (ATAGI) produced a helpful document on ‘Weighing up the potential benefits against risk of harm from COVID-19 vaccine AstraZeneca’ [11] to address concerns of vaccine-associated thrombosis with thrombocytopenia syndrome. While ATAGI released a clinical ‘Guidance on myocarditis and pericarditis after mRNA COVID-19 vaccines’ [12], there have not been any documents focused on risk-benefit analysis.

By October 2021, 23.4% and 55.1% of Australians aged over 16 years had received one and two doses of a COVID-19 vaccine, respectively, and an unspecified but small percentage had received a third dose [13]. Because of concerns related to the risk of thrombosis and thrombocytopenia syndrome with the AstraZeneca COVID-19 vaccine, the Pfizer vaccine was the standard recommendation for those aged <60 years [14]. However, six-month Pfizer vaccine effectiveness data that became available in October 2021 showed concerning reductions in protection against symptomatic infection each month after administration of the second dose [15]. In the context of the reopening of Australian borders in December 2021 and the introduction of the highly transmissible omicron variant, this decrease in vaccine effectiveness may leave even those who have had two doses of a COVID-19 vaccine at substantial risk of developing symptomatic COVID-19. Even for the highly vaccinated population of Australia, it was therefore crucial to communicate the necessity of third doses for maintaining optimal protection against symptomatic infection, serious illness, and death.

To effectively facilitate this communication, a risk-benefit analysis tool capable of integrating best evidence from multiple data sources (both Australian and international) and formats (government reports, published literature, and expert opinion) is required [16]. Furthermore, this tool must be easy to update as the pandemic landscape rapidly evolves and as more data become available. We have previously developed a Bayesian network (BN) model to analyze the risks and benefits of the COVID-19 AstraZeneca vaccine in the Australian population [17,18]. This model was used to program the COVID-19 Risk Calculator (CoRiCal) [19], a user-friendly online tool that enables scenario analysis based on user inputs (age, sex, vaccination status, transmission scenario). The tool provides probability estimates for targeted subgroups and can be used by health managers as well as individuals alone or in conjunction with their GP for shared decision making on vaccination. This study describes the BN model used to program the second version of the CoRiCal tool, and results of population-level risk-benefit analysis of the Pfizer COVID-19 vaccine for the Australian context.

## 2. RESULTS

### 2.1. Model description

The BN model was designed to predict five outcomes:

i. Probability of developing and dying from Pfizer vaccine-associated myocarditis (n5, n12) – depending on vaccine dose (n1), age (n2), and sex (n3);
ii. Background probability of developing and dying from myocarditis (in those who have not had Pfizer vaccine or COVID-19) (n6, n13). Estimates were converted to probability of events over two months to enable comparison with the probability of vaccine-associated (n5, n12) and infection-associated outcomes (n10, n14) over two-month periods;
iii. Probability of symptomatic COVID-19 (n10) – depending on intensity of community transmission (n4), vaccine effectiveness against symptomatic infection (n7), relative risk of symptomatic infection by age and sex (n9);
iv. Probability of dying from COVID-19 (n14) – depending on age (n2), sex (n3), vaccine effectiveness against death (n8); and
v. Probability of developing and dying from COVID-19-related myocarditis (n11, n15) – depending on age (n2), sex (n3).

The BN (Figure 1) displays the links between variables and outcomes based on the assumptions presented in Table 1 [11,15,20–35] and Supplementary Tables S1-9. Table 2 summarises each of the 15 nodes and their parent/child associations.

**Table 1.**
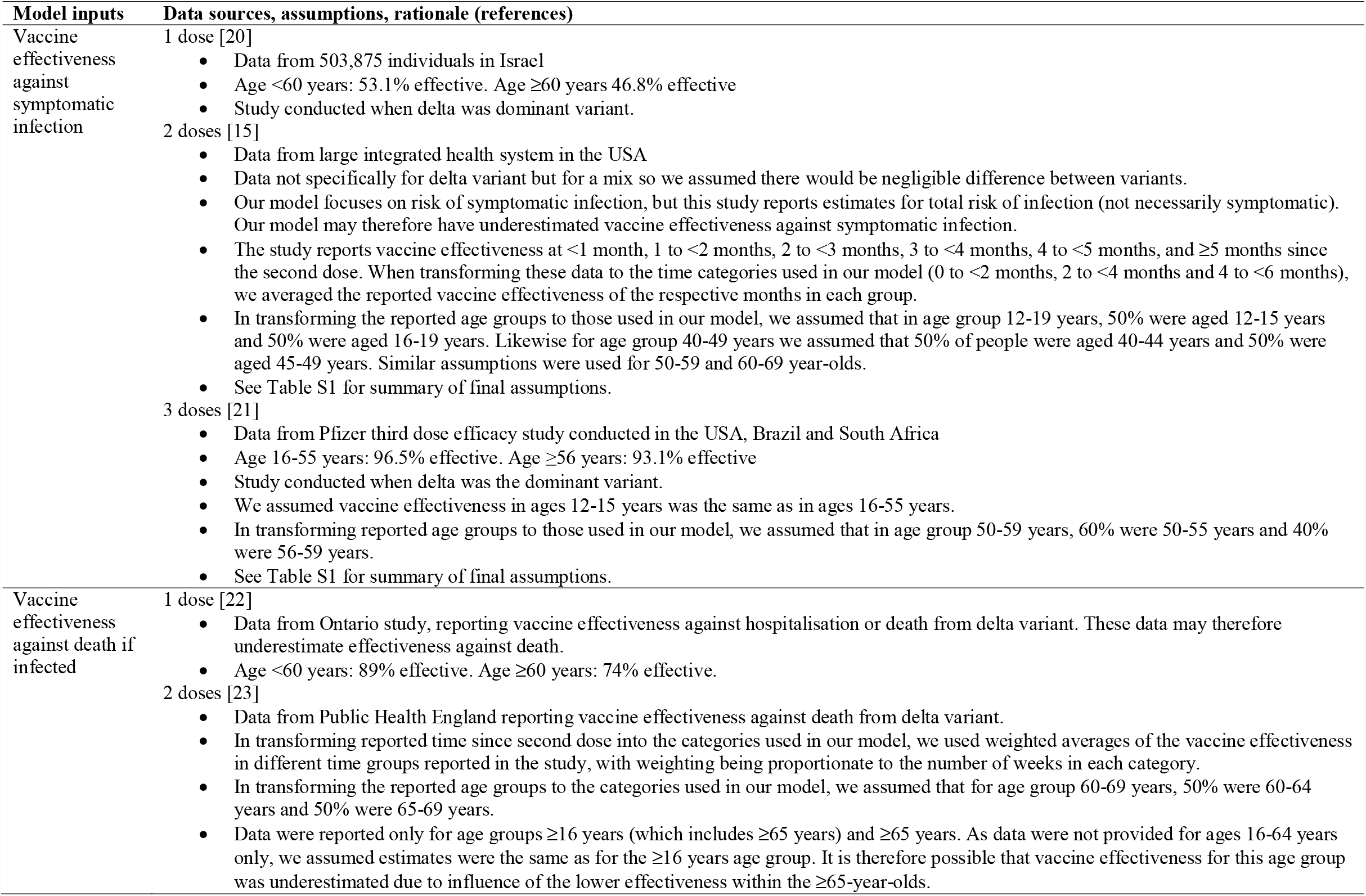
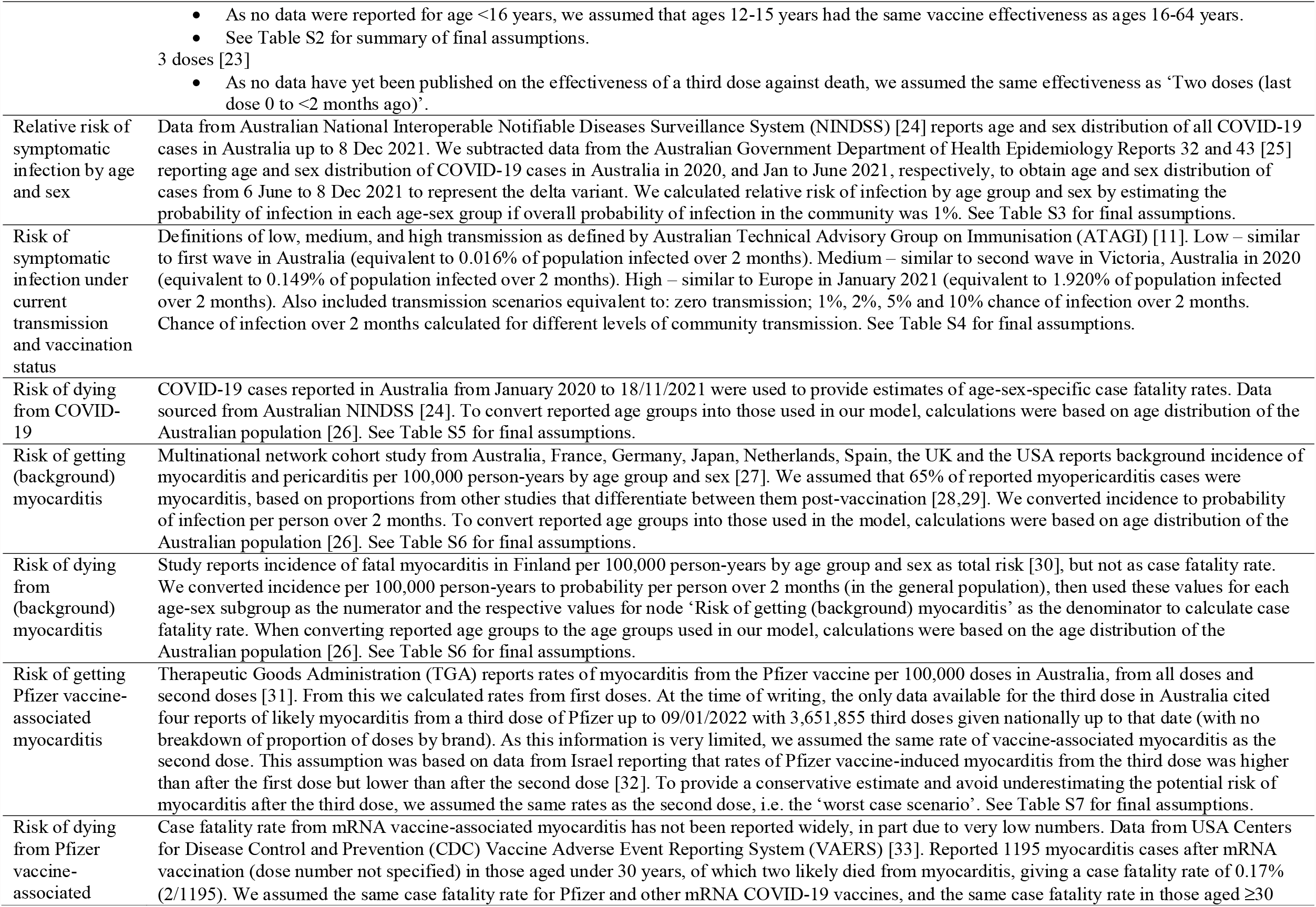
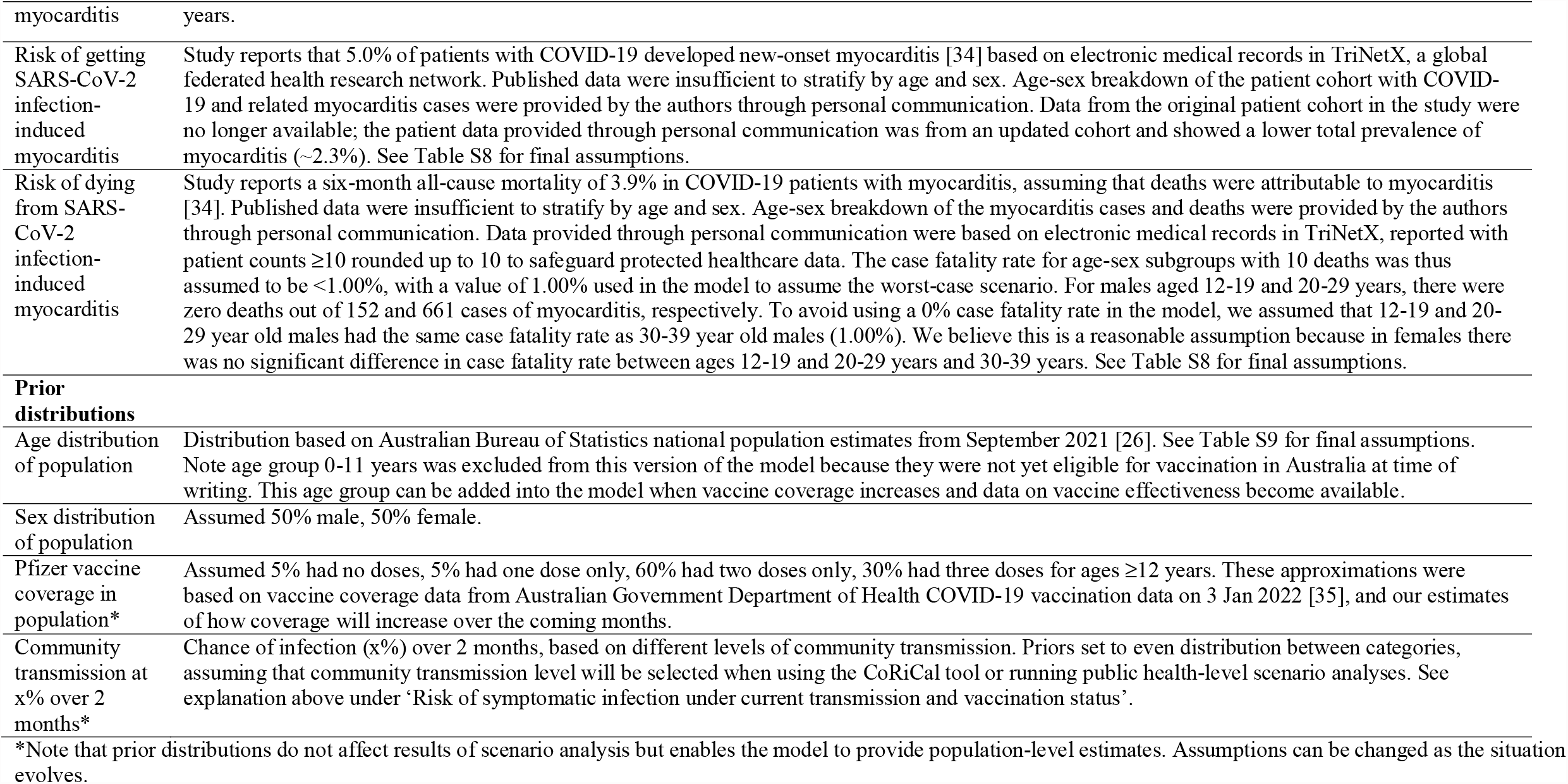
Summary of data sources, assumptions, and prior distributions for a Bayesian network to assess risks versus benefits of the Pfizer COVID-19 vaccine.

**Table 2.** Summary of nodes and relationships between nodes in a Bayesian network for assessing risks versus benefits of the Pfizer COVID-19 vaccine.

| Node name (number) | Description | Potential values | Node type | Parent nodes | Child nodes |
| --- | --- | --- | --- | --- | --- |
| Pfizer vaccine dose & time since dose 2 (n1) | Vaccine dose number | None,<br>1<sup>st</sup> dose (<3 weeks ago),<br>2<sup>nd</sup> dose (last dose 0 to <2 months ago),<br>2<sup>nd</sup> dose (last dose 2 to <4 months ago),<br>2<sup>nd</sup> dose (last dose 4 to <6 months ago),<br>3<sup>rd</sup> dose | Input | Age group (n2) | n5, n7, n8 |
| Age group (n2) | Age group (years) | 12-19, 20-29,30-39, 40-49, 50-59, 60-69, ≥70 | Input | N/A – Default priors: population distribution of Australia by age | n1, n5-9, n11, n13-15 |
| Sex (n3) | Sex | Male, female | Input | N/A – Defaults to uniform distribution | n5, n6, n9, n11, n13-15 |
| Community transmission at x% over 2 months (n4) | Probability of infection over 2 months based on different levels of community transmission | None, ATAGI definitions of low, med, high, 1%, 2%, 5%, 10% | Input | N/A – Defaults set to uniform distribution | n10 |
| Vaccine-associated myocarditis (n5) | Probability of developing myocarditis from the Pfizer COVID-19 vaccine | Yes, no | Intermediate | Pfizer vaccine dose & time since dose 2 (n1), Age group (n2), Sex (n3) | n12 |
| Background myocarditis over 2 months (n6) | Probability of developing myocarditis over 2 months (background rate in those who have not had vaccine or infection) | Yes, no | Outcome | Age group (n2), Sex (n3) | n13 |
| Vaccine effectiveness against symptomatic infection (n7) | Effectiveness of the vaccine at preventing symptomatic SARS-CoV-2 infection | Effective, ineffective | Intermediate | Pfizer vaccine dose & time since dose 2 (n1), Age group (n2) | n10 |
| Vaccine effectiveness against death (n8) | Effectiveness of the vaccine at preventing deaths from symptomatic SARS-CoV-2 infection | Effective, ineffective | Intermediate | Pfizer vaccine dose & time since dose 2 (n1), Age group (n2) | n14 |
| Relative risk of symptomatic infection by age and sex (n9) | Relative risk of symptomatic SARS-CoV-2 infection depending on age and sex | Yes, no | Intermediate | Age group (n2), Sex (n3) | n10 |
| Risk of symptomatic infection under current transmission and vaccination status (n10) | Probability of symptomatic COVID-19 | Yes, no | Intermediate | Community transmission at x% over 2 months (n4), Vaccine effectiveness against symptomatic infection (n7), Relative risk of symptomatic infection by age and sex (n9) | n11, n14 |
| Myocarditis from COVID-19 (n11) | Probability of developing myocarditis related to SARS-CoV-2 infection | Yes, no | Intermediate | Age group (n2), Sex (n3), Risk of symptomatic infection under current transmission and vaccination status (n10) | n15 |
| Die from vaccine-associated myocarditis (n12) | Probability of dying from COVID-19 vaccine-associated myocarditis | Yes, no | Outcome | Vaccine-associated myocarditis (n5) | N/A |
| Die from myocarditis (background) (n13) | Probability of dying from myocarditis (background rate in those who have not had COVID-19 vaccine or SARS-CoV-2 infection) | Yes, no | Outcome | Age group (n2), Sex (n3), Background myocarditis over 2 months (n6) | N/A |
| Die from COVID-19 (n14) | Probability of dying from COVID-19 | Yes, no | Outcome | Age group (n2), Sex (n3), Vaccine effectiveness against death (n8), Risk of symptomatic infection under current transmission and vaccination status (n10) | N/A |
| Die from COVID-19-related myocarditis (n15) | Probability of dying from COVID-19-related myocarditis | Yes, no | Outcome | Age group (n2), Sex (n3), Myocarditis from COVID-19 (n11) | N/A |

ATAGI: Australian Technical Advisory Group on Immunisation.

**Figure 1.**
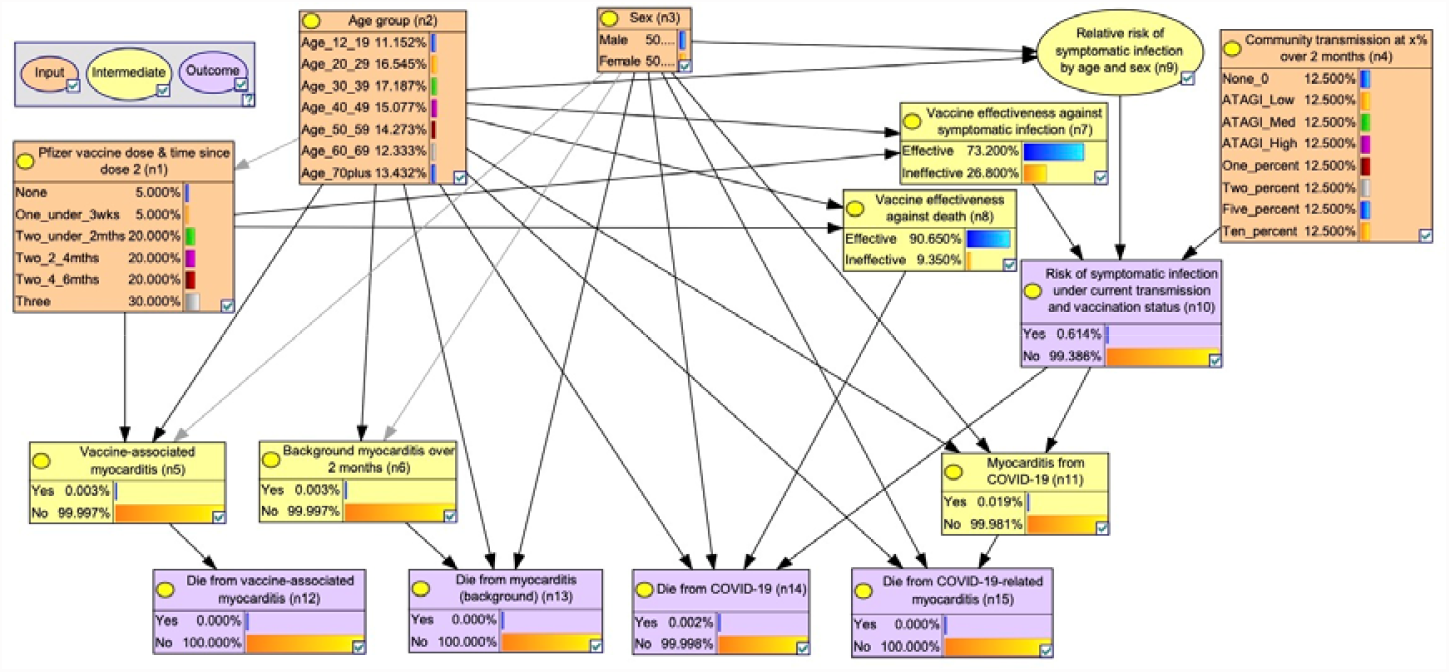
Bayesian network for assessing risks versus benefits of the Pfizer COVID-19 vaccine in Australia, with nodes in their default states.

The BN includes four input nodes (orange) for use in scenario analyses: Pfizer vaccine dose and time since second dose (n1), age (n2), sex (n3), and intensity of community transmission (n4). Community transmission scenarios were presented as probability of infection over two months to enable comparison of vaccination risks versus benefits, as vaccine effectiveness is expected to decrease over time (modelled using two-month intervals for time since second dose). Transmission scenarios were based on ATAGI definitions of low/medium/high risk [11] (equivalent to x, y, z% chance of infection over two months), and 1%, 2%, 5% and 10% chance of infection over two months. The model contains six intermediate nodes (yellow): Pfizer vaccine-associated myocarditis (n5), background incidence of myocarditis (n6), vaccine effectiveness (n7, n8), relative risk of symptomatic infection based on age and sex (n9), and incidence of COVID-19-related myocarditis (n11).

Two model versions were constructed employing distinct definitions of the ‘Pfizer vaccine dose and time since dose 2’ node (n1):

- Version 1: Pfizer vaccine doses defined as no doses, first dose, second dose, and third dose. This version allows estimation of the probability of vaccine-associated myocarditis with each dose of vaccine.
- Version 2: Pfizer vaccine doses defined as no doses, received only one dose, received two doses, and received three doses. This version allows estimation of the probability of deaths in the target population based on vaccine coverage rates.

### 2.2. Model validation

All authors agreed that the final model accurately represented the variables, their states, and associations within the model’s scope, in a manner consistent with the best current evidence. Model predictions were matched by independent calculations of selected outcome probabilities (Supplementary Table S10).

### 2.3. Risk-benefit analysis

#### 2.3.1. Estimated risks of background myocarditis, Pfizer vaccine-associated myocarditis and myocarditis in patients with symptomatic COVID-19

Based on background rates of myocarditis reported by Li et al. [27] and Barda et al. [28], our model estimated two-month incidence of 10.0 (females aged 12-19 years) to 53.9 (males aged ≥70 years) cases per million, and overall case fatality rate (CFR) ranging from 1.2% to 4.3% for different age-sex subgroups (Supplementary Table S6).

Up to 09/12/2021 in Australia, age-sex-specific incidence of Pfizer vaccine-associated myocarditis cases ranged from zero to 24 per million after the first dose, and zero to 103 per million after the second dose (Supplementary Table S7), with no reported deaths. Our model assumed an overall CFR of 0.17% (two deaths out of 1195 cases) based on reports from the Centers for Disease Control and Prevention Vaccine Adverse Event Reporting System in the USA [33] (Table 1).

At the time of writing, Australian data on myocarditis in COVID-19 patients were limited (Table 1). Model assumptions on the incidence and CFR of myocarditis in COVID-19 patients were obtained from an international cohort study by Buckley et al. [34], and additional unpublished age-sex specific data from the study via personal communication with the lead author. Data showed incidence ranging from 1.66% to 13.74%, and CFR ranging from <1% to 15.14%, depending on age and sex (Supplementary Table S8). Based on estimates from model version 2, Figure 2 shows that, in a population aged ≥12 years, with vaccine coverage of 5% unvaccinated, 5% had one dose, 60% had two doses and 30% had three doses, the probability of developing myocarditis related to symptomatic COVID-19 was 239 to 5847 times higher than developing Pfizer vaccine-associated myocarditis, depending on age group and sex (Figure 2, dashed lines). The probability of dying from myocarditis related to symptomatic COVID-19 was 1430 to 384,684 times higher than dying from vaccine-associated myocarditis, again depending on age group and sex (Figure 2, solid lines).

**Figure 2.**
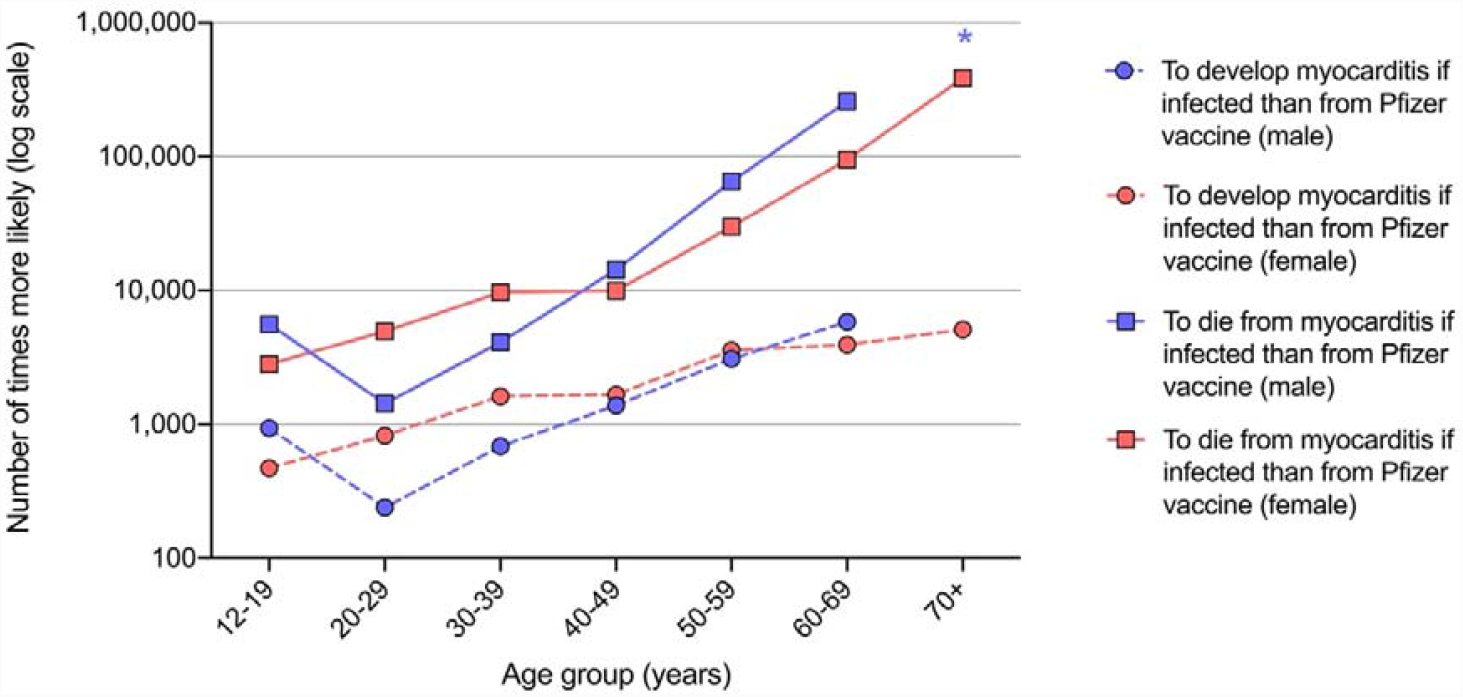
Number of times more likely (in log scale) to develop (circles) and die (squares) from myocarditis in patients with symptomatic COVID-19 than from Pfizer vaccine-associated myocarditis, by age group and sex. *For males aged ≥70 years, Pfizer vaccine-associated myocarditis had an incidence of 0%.

#### 2.3.2. Estimated symptomatic COVID-19 cases and deaths prevented

Model version 2 was used to calculate expected symptomatic COVID-19 cases and deaths prevented over two months per million population aged ≥12 years, where 5% were unvaccinated, 5% had one dose, 60% had two doses (20% each with the last dose administered 0 to <2, 2 to <4 and 4 to <6 months ago) and 30% had three doses. Figure 3a and 3b show the expected cases and deaths, respectively, prevented by age group under different community transmission intensities:

**Figure 3.**
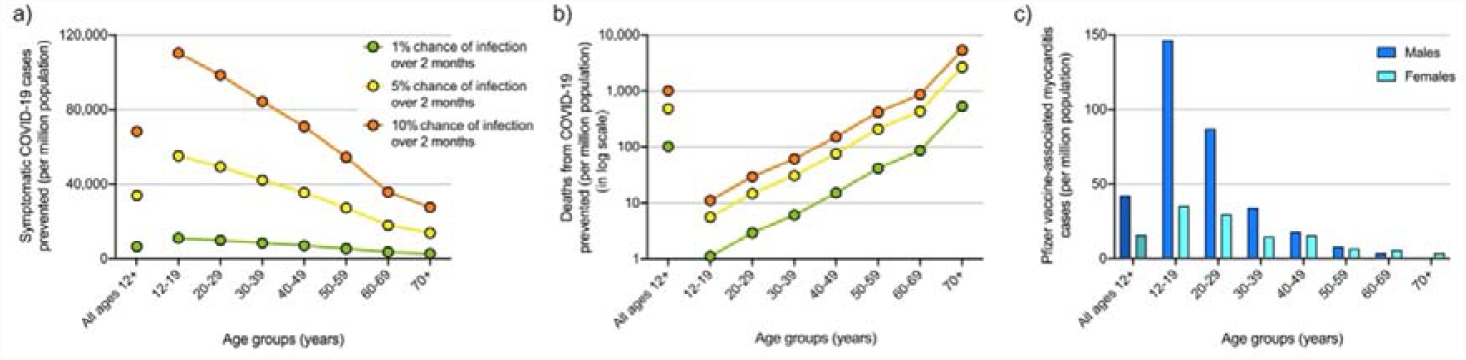
Estimated COVID-19 cases **(a)** and deaths **(b)** (in log scale) prevented over two months per million population of each age group if 5% had no doses, 5% had first dose, 60% had two doses (evenly distributed over 0 to <2, 2 to <4 and 4 to <6 months since second dose) and 30% had three doses of Pfizer COVID-19 vaccine if community transmission equivalent to 1% (green), 5% (yellow), and 10% (orange) chance of infection over two months. **(c)** Estimated cases of Pfizer COVID-19 vaccine-associated myocarditis over two months under the same vaccine coverage.

- 1% chance of infection over two months (green), equivalent to average of 3645 cases per day in Australia;
- 5% chance of infection over two months (yellow), equivalent to average of 7290 cases per day in Australia; and
- 10% chance of infection over two months (orange), equivalent to average of 18,225 cases per day in Australia.

The model estimates that for a million 12-19 year-olds with this vaccine coverage, 11,029 symptomatic COVID-19 cases and one death would be expected to be prevented under 1% transmission (green) versus 110,288 cases and 11 deaths prevented under 10% transmission (orange), with 146 expected cases of Pfizer vaccine-associated myocarditis in males and 35 cases in females (Figure 3b). In contrast, for a million people aged ≥70 years, 2757 cases and 98 deaths would be expected to be prevented under the 1% transmission scenario, 27,566 cases and 981 deaths prevented under the 10% transmission scenario, with less than five expected vaccine-associated myocarditis cases in males or females. Calculations are detailed in Supplementary Table S11.

#### 2.3.3. Estimated symptomatic COVID-19 cases and deaths under different vaccination coverage scenarios

Model version 2 was further used to estimate expected symptomatic COVID-19 cases and deaths per million people if transmission intensity was equivalent to a 10% chance of infection over two months, if 5% were unvaccinated, 5% had one dose, 60% had two doses and 30% had three doses (scenario one) (Figure 4, orange), versus if 0% of the population received no doses, 5% received the first dose only, 15% had two doses (5% each with the second dose administered 0 to <2, 2 to <4 and 4 to <6 months ago), and 80% had three doses (scenario two) (Figure 4, blue).

**Figure 4.**
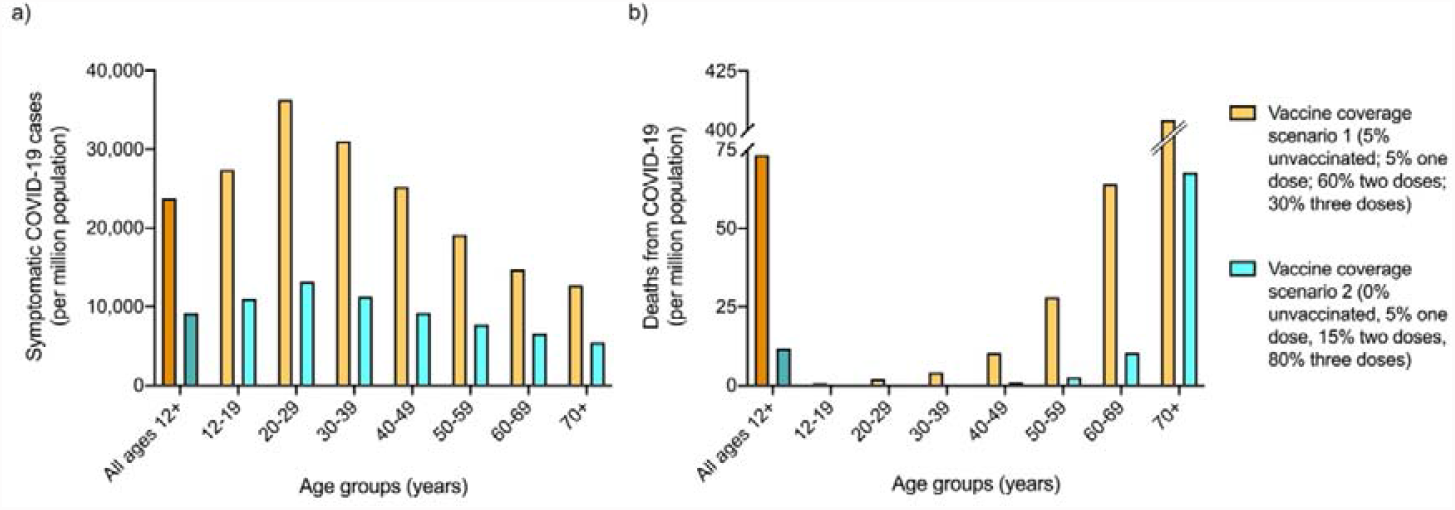
Comparison of expected number of COVID-19 cases **(a)** and deaths **(b)** per million population by age groups under vaccine coverage scenario one (5% had no doses, 5% had first dose, 60% had two doses [evenly distributed across time since second dose], and 30% had three doses of Pfizer COVID-19 vaccine), versus coverage scenario two (0% had no doses, 5% had one dose, 15% had two doses [evenly distributed across times since second dose] and 80% had three doses), under a transmission scenario equivalent to 10% chance of infection over two months.

The model shows that for a million people aged 12-19 years with the vaccine coverage described in scenario one, 27,391 symptomatic COVID-19 cases and less than one death from COVID-19 would be expected under 10% transmission over two months, versus 11,042 cases and less than one death in scenario two. For one million people aged 20-29 years, 36,249 cases and two deaths could be expected in scenario one versus 13,168 cases and less than one death under scenario two. In contrast, for a million people aged ≥70 years, 12,694 cases and 404 deaths would be expected in scenario one versus 5487 cases and 68 deaths under scenario two.

### 2.4. Sensitivity analysis

#### 2.4.1. Incidence of Pfizer vaccine-associated myocarditis

Therapeutic Goods Administration (TGA) reports between 14/10/2021 and 09/12/2021 [36] presented slight fluctuations in Pfizer vaccine-associated myocarditis incidence in Australia ranging from two to 37 cases per million depending on age-sex subgroup (Table 3). These small changes exerted no substantive impact on population-level estimates of the number of deaths. Model calculations also showed expected Pfizer vaccine-associated myocarditis deaths per million second doses to change only slightly during this time; differences ranged from 0.000 to 0.063 deaths per million by age-sex subgroup when comparing data from 14/10/2021 and 09/12/2021.

**Table 3.** Evolving evidence on incidence of Pfizer vaccine-associated myocarditis by age and sex in Australia in October-December 2021.

| Age group (years) | Estimated incidence of myocarditis per million 2<sup>nd</sup> doses<sup>a</sup> |  |  |  | Estimated deaths per million 2<sup>nd</sup> doses based on 0.34% CFR<sup>b</sup> |  |  |  | Difference in estimated cases per million 2<sup>nd</sup> doses compared to 14/10/21 |  | Difference in estimated deaths per million 2<sup>nd</sup> doses compared to 14/10/21 |  |
| --- | --- | --- | --- | --- | --- | --- | --- | --- | --- | --- | --- | --- |
|  | 14/10/21 |  | 09/12/21 |  | 14/10/21 |  | 09/12/21 |  | 09/12/21 |  | 09/12/21 |  |
|  | Male | Female | Male | Female | Male | Female | Male | Female | Male | Female | Male | Female |
| **12-19** | 75 | 14 | 103 | 25 | 0.128 | 0.024 | 0.175 | 0.043 | 28 | 11 | 0.048 | 0.019 |
| **20-29** | 22 | 12 | 59 | 19 | 0.037 | 0.020 | 0.100 | 0.032 | 37 | 7 | 0.063 | 0.012 |
| **30-39** | 6 | 3 | 15 | 6 | 0.010 | 0.005 | 0.026 | 0.010 | 9 | 3 | 0.015 | 0.005 |
| **40-49** | 10 | 10 | 11 | 9 | 0.017 | 0.017 | 0.019 | 0.015 | 1 | 1 | 0.002 | 0.002 |
| **50-59** | 3 | 3 | 1 | 4 | 0.005 | 0.005 | 0.002 | 0.007 | 2 | 1 | 0.003 | 0.002 |
| **60-69** | 0 | 0 | 0 | 0 | 0 | 0 | 0 | 0 | 0 | 0 | 0 | 0 |
| **≥70** | 0 | 0 | 0 | 0 | 0 | 0 | 0 | 0 | 0 | 0 | 0 | 0 |

<sup>a</sup>Incidence of myocarditis in Australia reported by Therapeutic Goods Administration (TGA). [31]
<sup>b</sup>CFR: Case fatality rate for all ages combined, calculated to be 0.17%, from [33].

#### 2.4.2. Vaccine effectiveness against symptomatic COVID-19 infection and death

The model calculated that in a population where 5% are unvaccinated, 5% had one dose, 60% had two doses and 30% had three doses, a hypothetical 5% or 10% decrease in vaccine effectiveness against the delta variant would result in a 17.8% or 35.7% increase in estimated symptomatic cases, respectively, and a 23.9% or 54.7% increase in estimated expected deaths, respectively (Table 4). Thus, model estimates of cases and deaths are highly sensitive to reductions in vaccine effectiveness, necessitating frequent monitoring of and updating with emerging vaccine effectiveness data, particularly against new variants.

**Table 4.** Impact of theoretical reduction in vaccine effectiveness against delta variant on estimated deaths, assuming 5% of population is unvaccinated, 5% had one dose, 60% had two doses and 30% had three doses.

|  | **Current**<br>**model**<br>**assumptions** | **If 5% less**<br>**effective** | **If 10% less**<br>**effective** |
| --- | --- | --- | --- |
| **Average vaccine effectiveness for all ages ≥12**<br>**against symptomatic infection after** |  |  |  |
| 1<sup>st</sup> dose (<3 weeks ago) | 51.5% | 46.5% | 41.5% |
| 2<sup>nd</sup> dose (last dose 0-<2 months ago) | 85.3% | 80.3% | 75.3% |
| 2<sup>nd</sup> dose (last dose 2-<4 months ago) | 72.1% | 67.1% | 62.1% |
| 2<sup>nd</sup> dose (last dose 4-<6 months ago) | 52.6% | 47.6% | 42.6% |
| 3<sup>rd</sup> dose | 95.4% | 90.4% | 85.4% |
| **% Increase in estimated symptomatic cases**<br>**compared to current model assumptions of**<br>**vaccine effectiveness** | N/A | 17.7% | 35.4% |
| **Average vaccine effectiveness for all ages ≥12**<br>**against death after** |  |  |  |
| 1<sup>st</sup> dose (<3 weeks ago) | 85.1% | 80.1% | 75.1% |
| 2<sup>nd</sup> dose (last dose 0-<2 months ago) | 98.0% | 93.0% | 88.0% |
| 2<sup>nd</sup> dose (last dose 2-<4 months ago) | 95.2% | 90.2% | 85.2% |
| 2<sup>nd</sup> dose (last dose 4-<6 months ago) | 91.8% | 86.8% | 81.8% |
| 3<sup>rd</sup> dose | 98.0% | 93.0% | 88.0% |
| **% Increase in estimated deaths compared to**<br>**current model assumptions of vaccine**<br>**effectiveness** | N/A | 23.8% | 54.9% |

## 3. DISCUSSION

We developed a BN model to facilitate risk-benefit analysis of the Pfizer COVID-19 vaccine for the Australian population. Results from this model highlight the importance of both individual factors such as age, sex, and vaccination status, and location-specific factors that reflect the current pandemic landscape, such as transmission intensity, case incidence and CFR from COVID-19, and COVID-19- and Pfizer vaccine-associated myocarditis. Our model could be used to help inform discussions and decision-making for population health managers, individuals and clinicians. In this way, the model may aid in policy development, public health management, increased public awareness and improved shared-decision-making in medical consultations.

For Australians ≥12 years, we compared the risk of developing Pfizer vaccine-associated myocarditis, with the benefit of protection against developing and dying from symptomatic COVID-19 over two months under different transmission scenarios, if 5% were unvaccinated, 5% had a first dose, 60% had two doses, and 30% had three doses. Overall, an Australian is 471 to 5847 times more likely to develop COVID-19-related than vaccine-associated myocarditis, and 1430 to 384,684 times more likely to die from it, depending on age and sex (Figure 2). Under any transmission level, younger age groups benefited the most from protection against symptomatic COVID-19 while older age groups benefited the most from protection against fatal COVID-19 (Figure 3). Younger age groups were at higher risk of developing vaccine-associated myocarditis than older groups, and males were at greater risk than females. We note that myocarditis was more common after COVID-19 compared to the background rates, especially in younger men. In comparison, vaccine-associated myocarditis also has a predilection for younger males but at a much lower prevalence than cases associated with symptomatic COVID-19. Importantly, in the main, vaccination is justified in all age groups because myocarditis is generally mild in the young [37–39], and there is unequivocal evidence for reduced mortality in older individuals across all levels of community transmission.

While the above risk-benefit analyses were conducted assuming the Australian vaccine coverage at the time of writing, outcomes under other coverage rates can be assessed by the model. We compared the number of COVID-19 cases and deaths expected if the chance of infection was 10% over two months under a scenario where 5% are unvaccinated, 5% had a first dose, 60% had two doses and 30% had three doses, to those expected under a second scenario where 0% are unvaccinated, 5% had a first dose, 15% had two doses and 80% had three doses (Figure 4). Younger age groups benefited from the steepest decline in expected case rates, with at least 23,000 fewer cases per million in 20-29 year-olds. In contrast, older age groups benefited from the greatest decrease in expected deaths from COVID-19, with 337 fewer deaths per million expected in those aged ≥70 years.

Sensitivity analysis showed model estimates to be robust against minor changes in the number of Pfizer vaccine-associated myocarditis cases (Table 3), but highly affected by changes in vaccine effectiveness against symptomatic infection and death (Table 4). At a public health level, this holds important implications for COVID-19 burden if new variants such as omicron, for which vaccine effectiveness is decreased, continue to emerge or if vaccine effectiveness proves to wane over time. While vaccine effectiveness would have to drop to a very low threshold for the associated myocarditis risk to outweigh the benefit of protection against symptomatic infection and death from COVID-19 in any age-sex-subgroup, this result highlights the importance of updating the model as new evidence becomes available, or new variants emerge.

Model estimates must be contextualised within the scope of the BN model, which does not currently consider comorbidities or personal behaviour that may influence an individual’s risks of acquiring COVID-19, their response to the infection, or their individual risk of myocarditis. Furthermore, limitations to the availability of Australian data introduces uncertainty in the model inputs, so results may change as more data become available. For example, at the time of writing no Australian data were available on the incidence of Pfizer vaccine-associated myocarditis after the third dose and international data were deemed inappropriate as a substitute (see Table 1 assumptions), necessitating the use of rates for the second dose as a worst-case scenario. In another example, when calculating the delta variant-specific CFR from COVID-19, ideally CFR for the unvaccinated population would be used, and the 2-3 week lag between diagnosis and death accounted for. This information was not available in Australia, so the assumptions were made that the time-window of a few months for the delta wave was long enough to minimise the effect of time lag from infection to death, and the great majority of deaths during the delta wave was in unvaccinated people. Other limitations arise from the model development process, where the use of expert elicitation may be perceived to introduce bias in the evidence viewed. This was minimised through broad literature searches and frequent meetings with external experts such as cardiologists about the quality of the data sources used in the model assumptions.

Despite these limitations, the use of an evidence-based BN to model the risks and benefits of COVID-19 vaccination has many advantages. BNs allow for interactive scenario analysis so the model was well-suited for use in programming CoRiCal, a free online tool aimed at better informing the public and helping clinicians to best advise patients on the risks and benefits of COVID-19 vaccination [19]. Another benefit of BNs is the ease of updating, allowing for future model updates to incorporate other outcomes such as long COVID, different patient groups such as those <12 years and those with comorbidities, other vaccines such as Moderna, or different vaccine adverse events such as anaphylaxis. Finally, BNs are advantageous due to their integration of new data and different data sources in informing different aspects of the model. While this model has been designed for the Australian context, conditional probability tables (CPTs) can easily be re-populated wherever possible using data from another country.

In summary, we developed a BN to compare the risks and benefits of Pfizer COVID-19 vaccination in the Australian population in order to assist clinicians with providing guidance about the Pfizer COVID-19 vaccine. In a community rather than individual context, the final model can also be used to calculate population-level estimates to help inform policy development and public health management. Although designed to compare risks of developing and dying from COVID-19, COVID-19- and Pfizer vaccine-associated myocarditis for the delta variant, the model can be updated to consider the omicron or other variants, other inputs such as patient comorbidities, and other outcomes such as long COVID.

## 4. MATERIALS AND METHODS

### 4.1. Bayesian networks

BNs are graphical displays of directional associations between variables, as defined by conditional probabilities [40]. Nodes represent variables and have multiple potential states (e.g., male and female), and associations are represented by arrows in the direction of parent (independent) to child (dependent) variable (Figure 5). Probabilities are assigned to each potential node state via CPTs depending on parent node states or, in the case of no parents, prior distributions. The use of CPTs allows for integration of multiple data sources and formats including published figures, other literature and expert opinion, as well as easy updating when new data are presented [41]. BNs are also appropriate for analysing estimated or uncertain risks as they allow for sensitivity analysis to test multiple possible inputs [41].

**Figure 5.**
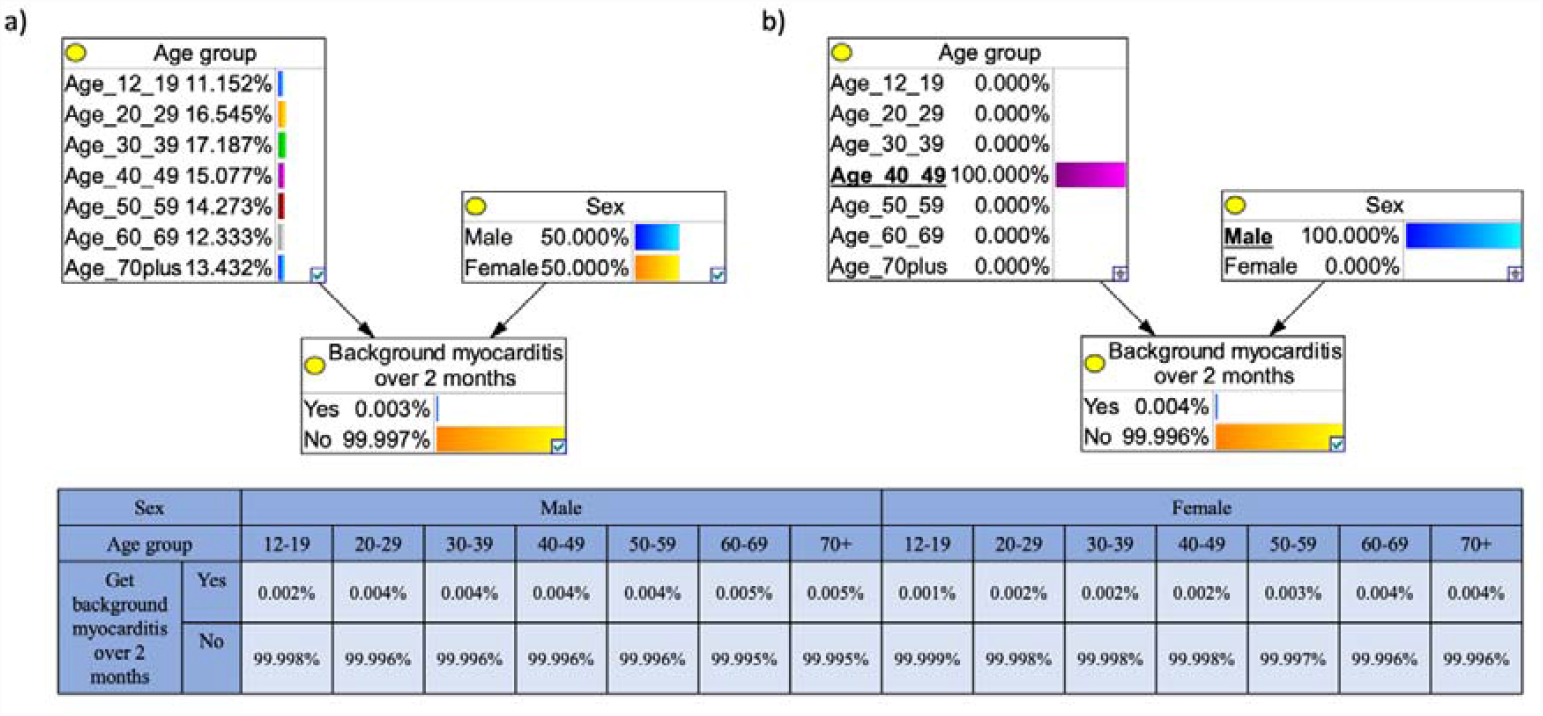
Example Bayesian network (BN) for modelling the risk of developing background myocarditis over 2 months based on age and sex. The output node, ‘Background myocarditis over 2 months’ is the child of two linked (black arrow) parent nodes, ‘Age group’, and ‘Sex’. As these parent nodes do not have parent themselves, the probabilities of each of their possible states are determined by a prior distribution; the model adopts the age distribution of the Australian population and an even distribution of males and females. The conditional probability table for the outcome node ‘Background myocarditis over 2 months’, gives the probability for each state of this node dependent on the parent node states. **(a)** In the default state, the BN shows that the chance of developing background myocarditis (not from COVID-19 or the Pfizer vaccine) over 2 months is 0.003% (e.g., in a population of 100,000 people, we expect three to get myocarditis in a two-month period). **(b)** An example of scenario analysis showing the chance of a 40-49 year old male (underlined) developing background myocarditis over two months, the model calculates a 0.004% chance of myocarditis.

Throughout the COVID-19 pandemic, BNs have been used in decision making [42], risk assessment [43] and analysis [44,45]. We have previously developed the first BN model for risk–benefit analysis of a COVID-19 vaccine, and used the model outputs to design an online tool to communicate the risks and benefits of the AstraZeneca COVID-19 vaccine in the Australian context [17–19].

### 4.2. Model design

The model was based on best evidence from multiple sources, designed through collaboration between subject matter experts (KRS, RP, JL, JES, SJB) and modelers (CLL, HJM, KM, JES) as described previously [17,18]. The model focuses on ages ≥12 years due to insufficient data on younger age groups at the time of development.

### 4.3. Myocarditis

Acute myocarditis can result in myocardial inflammation from either an infectious or immune-mediated aetiology [46]. Thus, our model compared the risk of Pfizer vaccine-associated myocarditis with the risk of myocarditis in COVID-19 patients. While often asymptomatic, myocarditis may present as chest pain, palpitations and/or dysrhythmias [46–48] and can cause dilated cardiomyopathy, arrhythmia and/or sudden cardiac death [47,48]. In Australia, myocarditis is often diagnosed using electrocardiogram, serum troponin levels, inflammatory markers, chest X-ray, echocardiography and occasionally endomyocardial biopsy [12]. However, these methods can underestimate the presence of myocarditis in comparison to more sensitive cardiac magnetic resonance imaging (MRI), which is considered the gold-standard for non-invasive diagnosis worldwide [49,50]. The 2018 Lake Louise criteria for MRI-based diagnosis of myocarditis targets tissue-based imaging markers of oedema, hyperaemia, necrosis and fibrosis [51,52]. To ensure the diagnosis of myocarditis was made robustly in our model, data reporting myocarditis cases diagnosed via cardiac MRI were used wherever possible.

While both the Pfizer COVID-19 vaccine and COVID-19 itself may also be associated with pericarditis, either separately or simultaneously with myocarditis, this model focuses solely on myocarditis. This is because diagnostic criteria for pericarditis are not well-defined, and because it is less common than myocarditis. In studies that reported ‘myocarditis/pericarditis’, we estimated that ∼65% of cases were attributable to myocarditis, based on proportions of cases reported in studies that differentiate between them [28,29].

The definitions for vaccine-associated and infection-induced myocarditis used for the model reflect those used within the studies from which data were drawn. Vaccine-associated myocarditis was defined as confirmed myocarditis within approximately 10 days of vaccine administration [31], and COVID-19-related myocarditis was defined as myocarditis that occurred within 6 months of COVID-19 diagnosis [34].

### 4.4. Data sources

CPTs were derived from data compiled by experts from published material, government reports, and through dialog with external clinical experts (e.g., cardiologists regarding the evidence for Pfizer vaccine-associated, COVID-19-related and background rate of myocarditis). Official Australian authority-issued data were employed whenever possible (e.g., national data on Pfizer vaccine-associated myocarditis). When this was unavailable, data were retrieved from other reliable and publicly available sources (e.g., background rates of myocarditis). Where Australian data were not readily available and international data were not suitable to use for the Australian context, expert opinion was sought. For example, there were limited data in Australia about Pfizer vaccine-associated myocarditis incidence and CFR after the third dose. While rates were reported in Israel and Singapore, these were deemed inappropriate to use in the model as reported rates from first and second doses in these countries were much lower than in Australia. However, both reported lower incidence of myocarditis after the third dose than the second dose. Therefore, to avoid underestimating the risk, the decision was made by the subject experts to use a conservative assumption that incidence after the third dose was the same as the second dose. For some variables, data analysis was required to obtain probabilities for the CPTs, e.g., converting COVID-19 case incidence into probability of infection over two months for the community transmission intensity node, or averaging data to fit the BN age categories. Table 1 and Supplementary Tables S1-9 summarise data sources, model assumptions, and rationale.

The BN incorporates default prior distributions for age group (based on the Australian population’s age distribution), sex (50% male, 50% female), and vaccine coverage (5% of the population unvaccinated, 5% of received one dose, 60% received two doses [20% with the second dose administered 0 to <2 months ago, 20% 2 to <4 months ago, and 20% 4 to <6 months ago], and 30% received three doses [administered approximately 3 weeks ago]). Prior distributions do not influence scenario analyses results, e.g., once male sex is selected, outputs relate only to males regardless of the entered prior distribution of sexes. Prior distributions can also be altered to model specific scenarios, e.g., different levels of vaccine coverage.

### 4.5. Model validation

Subject experts and modellers reviewed the final model to evaluate if the network structure, variables, relationships, and assumptions adequately portrayed the current best evidence. Multiple scenarios were defined, and model outputs manually calculated from the data sources and pre-defined assumptions to validate the BN’s predictive behaviour (Supplementary Table S10).

### 4.6. Risk-benefit analysis

We assessed the risks versus benefits of the Pfizer vaccine if 5% of the population received no doses, 5% received the first dose only, 60% had two doses (20% each with the last dose administered 0 to <2, 2 to <4 and 4 to <6 months ago), and 30% had three doses within the last two months (third dose administered 4 to 6 months after second dose). We assumed the same vaccine coverage for all age groups. These priors were selected to represent predicted vaccination coverage at the time of writing. We compared the following risks (vaccine-associated myocarditis) and benefits (potential COVID-19 cases and deaths prevented) assuming the above vaccination coverage:

i. Estimated number of times more likely for a person with symptomatic COVID-19 to develop and die from COVID-19-related myocarditis, than for a person to develop and die from Pfizer vaccine-associated myocarditis.
ii. Estimated symptomatic COVID-19 cases and deaths prevented per million population if transmission intensity was equivalent to 1%, 5% or 10% chance of infection over two months, versus estimated cases of Pfizer vaccine-associated myocarditis.
iii. Estimated symptomatic COVID-19 cases and deaths per million if transmission intensity was equivalent to 10% chance of infection over two months, under the vaccination coverage scenario described above versus a possible future scenario where 0% of the population received no doses, 5% received the first dose only, 15% had two doses (5% each with the last dose administered 0 to <2, 2 to <4 and 4 to <6 months ago), and 80% had three doses.

### 4.7. Sensitivity analysis

Evidence informing many model inputs rapidly evolved throughout the model development process. We ran sensitivity analyses for two variables considered most likely to fluctuate over time, to evaluate the necessary frequency for updating model assumptions.

From October-December 2021, reported Pfizer vaccine-associated myocarditis incidence in Australia increased weekly but numbers remained very low. We assessed TGA reports from 14/10/2021 and 09/12/2021 [36] to evaluate how changes in data influenced model predictions of age-sex-specific myocarditis cases from the second vaccine dose, per million people. We also assessed model output sensitivity to hypothetical 5% and 10% decreases in vaccine effectiveness against both symptomatic infection and death for the delta variant.

## Data Availability

All data generated or analysed during this study are included in this article and its supplementary files.

## 7. ACKNOWLEDGMENTS

We thank Kim Sampson from Immunisation Coalition and Dr Andrew Baird (St Kilda Medical Group, Australia) for facilitating the collaboration between authors; Aapeli Vuorinen (Data Science Institute, Columbia University, USA) for programming the online CoRiCal tool; Dr Michael Waller (School of Public Health, The University of Queensland, Australia) for contributions to model validation; A/Prof Hassan Valley (Deakin University, Melbourne, Australia) for contributions to discussions about risk communication and data visualisation; A/Prof Sudhir Wahi, Director of Echocardiography and Senior Staff Cardiologist, Cardiac Society of Australia and New Zealand (CSANZ) Imaging Council for their feedback on myocarditis-related data; and Dr Benjamin J.R. Buckley (University of Liverpool and Liverpool Heart & Chest Hospital, Liverpool, UK) for providing data on age and sex subgrouping of COVID-19-related myocarditis cases and deaths of the cohort described in [34]. Our BN model was built using GeNIe Modeler (BayesFusion 2019), available free of charge for academic research and teaching use from https://www.bayesfusion.com.

## 9. AUTHOR CONTRIBUTIONS

Conception and design: JL, KRS, CLL, JES

Acquisition of data: JL, RP, JES, CLL, KRS

Analysis and interpretation: JES, CLL, JL, HJM, KM, SJB Drafting the article: JES, CLL, HJM

Revising article for important intellectual content: All authors Final approval of submitted version: All authors

