## Supplementary Material for "Quantifying the risks versus benefits of the Pfizer COVID-19 vaccine in Australia: a Bayesian network analysis"

### Supplementary Materials

#### Table S1. Pfizer COVID-19 vaccine effectiveness against symptomatic infection with SARS-CoV-2 delta variant, by age group.

| Age (years) | 1 dose[a] | 2 doses (last dose 0 to <2 months ago)[b] | 2 doses (last dose 2 to <4 months ago)[b] | 2 doses (last dose 4 to <6 months ago)[b] | 3 doses (<4 months post 3rd dose)[c] |
|---|---|---|---|---|---|
| 12-19 | 53.1% | 89.0% | 79.5% | 74.0% | 96.5% |
| 20-29 | 53.1% | 86.5% | 73.0% | 48.0% | 96.5% |
| 30-39 | 53.1% | 86.5% | 73.0% | 48.0% | 96.5% |
| 40-49 | 53.1% | 86.3% | 72.8% | 51.8% | 96.5% |
| 50-59 | 53.1% | 86.0% | 72.5% | 55.5% | 95.1% |
| 60-69 | 46.8% | 82.8% | 69.0% | 50.8% | 93.1% |
| ≥70 | 46.8% | 79.5% | 65.5% | 46.0% | 93.1% |

Sources:

[a]Chodick, G., Tene, L., Patalon, T., Gazit, S., Tov, A.B., Cohen, D., and Muhsen, K. (2021). Assessment of effectiveness of 1 dose of BNT162b2 vaccine for SARS-CoV-2 infection 13 to 24 days after immunization. *JAMA Network Open* 4(6):e2115985. https://doi.org/10.1001/jamanetworkopen.2021.15985. [1]

[b]Tartof, S.Y., Slezak, J.M., Fischer, H., Hong, V., Ackerson, B.K., Ranasinghe, O.N., et al. (2021). Effectiveness of mRNA BNT162b2 COVID-19 vaccine up to 6 months in a large integrated health system in the USA: a retrospective cohort study. *The Lancet* 398(10309):1407–1416. https://doi.org/10.1016/S0140-6736(21)02183-8. [2]

[c]Perez, J.L. (2021). Efficacy and safety of BNT162b2 booster – C4591031 2 month interim analysis. *Centers for Disease Control and Prevention*. Accessed 17 December 2021 from https://www.cdc.gov/vaccines/acip/meetings/downloads/slides-2021-11-19/02-COVID-Perez-508.pdf. [3]

#### Table S2. Pfizer COVID-19 vaccine effectiveness against death if infected with SARS-CoV-2 delta variant, by age group.

| Age (years) | 1 dose[a] | 2 doses (last dose 0 to <2 months ago)[b] | 2 doses (last dose 2 to <4 months ago)[b] | 2 doses (last dose 4 to <6 months ago)[b] | 3 doses (<2 months post 3rd dose)[b] |
|---|---|---|---|---|---|
| 12-59 | 89% | 98.2% | 95.3% | 91.7% | 98.2% |
| 60-69 | 74% | 97.6% | 95.2% | 92.0% | 97.6% |
| ≥70 | 74% | 97.0% | 95.2% | 92.2% | 97.0% |

Sources:

[a]Nasreen, S., Chung, H., He, S., Brown, K.A., Gubbay, J.B., Buchan, S.A., et al. (2021). Effectiveness of mRNA and ChAdOx1 COVID-19 vaccines against symptomatic SARS-CoV-2 infection and severe outcomes with variants of concern in Ontario. *medRxiv*. https://doi.org/10.1101/2021.06.28.21259420. [4]

[b]Andrews, N., Tessier, E., Stowe, J., Gower, C., Kirsebom, F., Simmons, R., et al. (2021). *medRxiv*. https://doi.org/10.1101/2021.09.15.21263583. [5]

#### Table S3. Relative probability of infection by age group and sex for SARS-CoV-2 delta variant (chance of infection in each age-sex group if overall probability of infection of 1%).

| Age (years) | Male | Female |
|---|---|---|
| 0-11 | 1.37% | 1.30% |
| 12-19 | 1.41% | 1.34% |
| 20-29 | 1.41% | 1.29% |
| 30-39 | 1.18% | 1.12% |
| 40-49 | 0.98% | 0.94% |
| 50-59 | 0.76% | 0.72% |
| 60-69 | 0.51% | 0.49% |
| ≥70 | 0.39% | 0.42% |
| **Overall** | **1.03%** | **0.97%** |

Sources:

Australian Government Department of Health. (2021). Coronavirus (COVID-19) case numbers and statistics – cases and deaths by age and sex. *Australian Government Department of Health.* Accessed 17 December 2021 from https://www.health.gov.au/news/health-alerts/novel-coronavirus-2019-ncov-health-alert/coronavirus-covid-19-case-numbers-and-statistics#covid19-summary-statistics. [6]

Australian Government Department of Health. (2021). Coronavirus disease 2019 (COVID-19) epidemiology reports, Australia, 2020-2021. *Australian Government Department of Health.* Accessed 17 December 2021 from https://www1.health.gov.au/internet/main/publishing.nsf/Content/novel_coronavirus_2019_ncov_weekly_epidemiology_reports_australia_2020.htm. [7]

**Table S4. Probability of infection (over 2 months) based on different intensities of community transmission.**

| Intensity of community transmission | Cases per 100,000 over 16 weeks* | Cases per million over 2 months | Estimated % of population infected over 2 months | Equivalent to cases/day in Australia[a] |
|---|---|---|---|---|
| Zero | 0 | 0 | 0.000% | 0 |
| Low* | 29 | 157 | 0.016% | 58 |
| Medium* | 275 | 1,490 | 0.149% | 543 |
| High* | 3,544 | 19,197 | 1.920% | 6998 |
| 1% chance of infection over 2 months | | 10,000 | 1.000% | 3645 |
| 2% chance of infection over 2 months | | 20,000 | 2.000% | 7290 |
| 5% chance of infection over 2 months | | 50,000 | 5.000% | 18,225 |
| 10% chance of infection over 2 months | | 10,0000 | 10.000% | 36,450 |

* Definitions of low, medium, and high transmission (cases per 100,000 over 16 weeks) as defined by [9]. Low: similar to first wave in Australia. Medium: similar to second wave in VIC. High: similar to Europe in January 2021.
[a]Based on Australian population of 21.87 million. [8]
Source:

Australian Technical Advisory Group on Immunisation. (2021). Weighing up the potential benefits and risk of harm from COVID-19 vaccine AstraZeneca. *Australian Government Department of Health.* Accessed 17 December 2021 from https://www.health.gov.au/sites/default/files/documents/2021/06/covid-19-vaccination-weighing-up-the-potential-benefits-against-risk-of-harm-from-covid-19-vaccine-astrazeneca_2.pdf. [9]

**Table S5. Cases, deaths, and case fatality rate of COVID-19 in Australia in ages ≥12 years by age and sex, 1/1/2020 to 18/11/2021.**

| | Male | | | Female | | |
|---|---|---|---|---|---|---|
| **Age (years)** | **Cases** | **Deaths** | **Case fatality rate** | **Cases** | **Deaths** | **Case fatality rate** |
| 12-19 | 11,934 | 1 | 0.01% | 11,286 | 1 | 0.01% |
| 20-29 | 20,066 | 6 | 0.03% | 18,755 | 3 | 0.02% |
| 30-39 | 17,324 | 12 | 0.07% | 16,104 | 7 | 0.04% |
| 40-49 | 12,277 | 28 | 0.23% | 11,452 | 12 | 0.10% |
| 50-59 | 9252 | 70 | 0.76% | 8837 | 39 | 0.44% |
| 60-69 | 5598 | 144 | 2.57% | 5375 | 58 | 1.08% |
| ≥70 | 5059 | 789 | 15.60% | 5786 | 752 | 13.00% |
| **Total** | **81,510** | **1,050** | **1.29%** | **77,595** | **872** | **1.12%** |

Sources:

Australian Government Department of Health. (2021). Coronavirus (COVID-19) case numbers and statistics – cases and deaths by age and sex. *Australian Government Department of Health.* Accessed 17 December 2021 from https://www.health.gov.au/news/health-alerts/novel-coronavirus-2019-ncov-health-alert/coronavirus-covid-19-case-numbers-and-statistics#covid19-summary-statistics. [6]
Australian Bureau of Statistics. (2021). National, state and territory population. *Australian Bureau of Statistics.* Accessed 15 December 2021 from https://www.abs.gov.au/statistics/people/population/national-state-and-territory-population/mar-2021/31010do002_202103.xls. [8]

**Table S6. Estimated background incidence and fatality of myocarditis over 2 months (in populations who have not received the Pfizer COVID-19 vaccine and have not been diagnosed with COVID-19).**

| | Incidence of myocarditis over 2 months (per million population)[b] | | Incidence of fatal myocarditis over 2 months (per million population)[c] | | Case fatality rates from myocarditis | |
|---|---|---|---|---|---|---|
| **Age (years)[a]** | **Male** | **Female** | **Male** | **Female** | **Male** | **Female** |
| 12-19 | 19.0 | 10.0 | 0.3 | 0.3 | 1.3% | 2.5% |
| 20-29 | 40.1 | 17.3 | 0.5 | 0.3 | 1.2% | 1.7% |
| 30-39 | 40.1 | 20.6 | 0.9 | 0.4 | 2.3% | 2.2% |
| 40-49 | 40.1 | 23.8 | 1.3 | 0.7 | 3.3% | 3.0% |
| 50-59 | 44.4 | 28.7 | 1.1 | 0.9 | 2.6% | 3.1% |
| 60-69 | 50.9 | 35.8 | 1.3 | 1.0 | 2.5% | 2.9% |
| ≥70 | 53.9 | 39.6 | 1.6 | 1.7 | 3.0% | 4.3% |

Sources:

[a]Australian Bureau of Statistics. (2021). National, state and territory population. *Australian Bureau of Statistics.* Accessed 15 December 2021 from https://www.abs.gov.au/statistics/people/population/national-state-and-territory-population/mar-2021/31010do002_202103.xls. [8]

[b]Li, X., Ostropolets, A., Makadia, R., Shoaibi, A., Rao, G., Sena, A.G., et al. (2021). Characterising the background incidence rates of adverse events of special interest COVID-19 vaccines in eight countries: multinational network cohort study. *The BMJ* 2021(373):n1435. https://doi.org/10.1101/2021.03.25.21254315. [10]

[c]Kytö, V., Saraste, A., Voipio-Pulkki, L., and Saukko, P. (2007). Incidence of fatal myocarditis: a population-based study in Finland. American Journal of Epidemiology 165(5):570–574. https://doi.org/10.1093/aje/kwk076. [11]

**Table S7. Rates of myocarditis cases per million Pfizer COVID-19 vaccine doses in Australia by age and sex.**

| | First dose | | Second dose | | Third dose[a] | |
|---|---|---|---|---|---|---|
| **Age (years)** | **Male** | **Female** | **Male** | **Female** | **Male** | **Female** |
| 12-19 | 24 | 6 | 103 | 25 | 103 | 25 |
| 20-29 | 17 | 7 | 59 | 19 | 59 | 19 |
| 30-39 | 17 | 8 | 15 | 6 | 15 | 6 |
| 40-49 | 5 | 5 | 11 | 9 | 11 | 9 |
| 50-59 | 7 | 2 | 1 | 4 | 1 | 4 |
| 60-69 | 4 | 6 | 0 | 0 | 0 | 0 |
| ≥70 | 0 | 4 | 0 | 0 | 0 | 0 |

[a]Assumed the same rates as after second dose because no data were available for rates after third dose.

Source:

Therapeutic Goods Administration. (2021). COVID-19 vaccine weekly safety report – 09-12-2021. *Australian Government Department of Health.* Accessed 17 December 2021 from https://www.tga.gov.au/periodic/covid-19-vaccine-weekly-safety-report-09-12-2021. [12]

**Table S8. COVID-19-related myocarditis cases, deaths, and case fatality rate in ages ≥12 years by age and sex, up to 6 months post-myocarditis diagnosis.**

| | Male | | | | | Female | | | | |
|---|---|---|---|---|---|---|---|---|---|---|
| Age (years) | COVID-19 cases | Myocarditis cases | Incidence | Deaths | Case fatality | COVID-19 cases | Myocarditis cases | Incidence | Deaths | Case fatality |
| 12-19 | 1106 | 152 | 13.74% | 0[a] | <1.00% | 12,291 | 204 | 1.66% | ≤10[b] | <1.00% |
| 20-29 | 31,758 | 661 | 2.08% | 0[a] | <1.00% | 54,404 | 1321 | 2.43% | ≤10[b] | <1.00% |
| 30-39 | 43,723 | 1025 | 2.34% | ≤10[b] | <1.00% | 76,988 | 1849 | 2.40% | ≤10[b] | <1.00% |
| 40-49 | 41,971 | 1044 | 2.49% | 18 | 1.72% | 65,273 | 1690 | 2.59% | ≤10[b] | <1.00% |
| 50-59 | 51,473 | 1242 | 2.41% | 44 | 3.54% | 68,627 | 1644 | 2.40% | 23 | 1.40% |
| 60-69 | 57,880 | 1286 | 2.22% | 95 | 7.39% | 65,223 | 1458 | 2.24% | 59 | 4.05% |
| ≥70 | 66,431 | 1314 | 1.98% | 199 | 15.14% | 74,800 | 1452 | 1.94% | 183 | 12.60% |
| **Total** | **294,342** | **6724** | **2.28%** | **366** | **5.44%** | **417,606** | **9618** | **2.30%** | **305** | **3.17%** |

[a]For males aged 12-19 and 20-29 years, there were zero deaths out of 152 and 661 cases of myocarditis, respectively. To avoid using a 0% case fatality rate in the model, we assumed that 12-19 and 20-29 year old males had the same case fatality rate as 30-39 year old males (1.00%).

[b]Patient counts of ≤10 were rounded up to 10 to safeguard protected healthcare data. Related case fatality rates were thus assumed to be <1.00%., with a value of 1.00% used in the model to assume the worst-case scenario.

Source:

Personal communication from authors regarding patient cohort described in: Buckley, B.J.R., et al. (2021). Prevalence and clinical outcomes of myocarditis and pericarditis in 718,365 COVID-19 patients. *European Journal of Clinical Investigation* 51(11):e13669. https://doi.org/10.1111/eci.13679. [13]

**Table S9. Age distribution of Australian population, March 2021.**

| Age (years) | Population | % of total population |
|---|---|---|
| 0-11 | 3,828,247 | 14.90% |
| 12-19 | 2,438,423 | 9.49% |
| 20-29 | 3,617,689 | 14.08% |
| 30-39 | 3,757,954 | 14.63% |
| 40-49 | 3,296,519 | 12.83% |
| 50-59 | 3,120,900 | 12.15% |
| 60-69 | 2,696,731 | 10.50% |
| ≥70 | 2,936,879 | 11.43% |
| **Total** | **25,693,342** | **100.00%** |

Source:

[a]Australian Bureau of Statistics. (2021). National, state and territory population. *Australian Bureau of Statistics.* Accessed 15 December 2021 from https://www.abs.gov.au/statistics/people/population/national-state-and-territory-population/mar-2021/31010do002_202103.xls. [8]

**Table S10. Results of manual calculations used to validate the mathematical assumptions used to parameterise the model. Values provided are from two independent modellers (blue, green) and model estimates (purple).**

| Questions | | Calculated estimate |
|---|---|---|
| **1.** For a 30-39 year-old male, what is the chance of symptomatic infection under ATAGI high transmission if: | a) Not vaccinated | 0.022701<br>0.022656<br>0.022656 |
| | b) Had one dose (administered <3 weeks ago) | 0.010647<br>0.010626<br>0.010626 |
| | c) Had two doses (last dose 0-<2 months ago) | 0.003065<br>0.003059<br>0.003059 |
| | d) Had two doses (last dose 2-<4 months ago) | 0.006129<br>0.006117<br>0.006117 |
| | e) Had two doses (last dose 4-<6 months ago) | 0.011804<br>0.011781<br>0.011781 |
| | f) Had three doses | 0.000795<br>0.000793<br>0.000793 |
| **2.** For a 30-39 year-old male with symptomatic COVID-19, what is the chance of dying from COVID-19 if: | a) Not vaccinated | 0.000693<br>0.000693<br>0.000693 |
| | b) Had one dose (administered <3 weeks ago) | 0.000076<br>0.000076<br>0.000076 |
| | c) Had two doses (last dose 0-<2 months ago) | 0.000012<br>0.000012<br>0.000012 |
| | d) Had two doses (last dose 2-<4 months ago) | 0.000033<br>0.000033<br>0.000033 |
| | e) Had two doses (last dose 4-<6 months ago) | 0.000057<br>0.000057<br>0.000057 |
| | f) Had three doses | 0.000012<br>0.000012<br>0.000012 |
| * **3.** Under ATAGI high transmission, for one million 50-59 year-old females, if 5% have had no vaccine doses, 5% have had 1 dose only, 60% have had 2 doses only and 30% have had 3 doses: | a) How many cases of vaccine-induced myocarditis would we expect? | 6.700000<br>6.700000<br>6.700000 |
| | b) How many vaccine-associated myocarditis-induced deaths would we expect? | 0.022919<br>0.022919<br>0.022919 |
| * **4.** For one million ≥70 year-old males, if 5% have had no vaccine doses, 5% have had 1 dose only, 60% have had 2 doses only and 30% have had 3 doses: | a) How many symptomatic cases would we expect over 2 months if there was ATAGI medium transmission during this time? | 183.0304<br>183.2208<br>183.2208 |
| | b) How many deaths from COVID-19 would we expect over 2 months if there was ATAGI medium transmission during this time? | 6.379250<br>6.389558<br>6.389558 |
| **5.** If a 60-69 year-old female was diagnosed with COVID-19: | a) What are her chances of developing COVID-19-related myocarditis? | 0.022354<br>0.022354<br>0.022354 |
| | b) What are her chances of dying from COVID-19-related myocarditis (before diagnosis)? | 0.000905<br>0.000905<br>0.000905 |

| | | |
|---|---|---|
| Additional population risk: | c) What are her background chances of developing myocarditis? | 0.000036<br>0.000036<br>0.000036 |
| | d) What are her background chances of dying from myocarditis? | 0.000001<br>0.000001<br>0.000001 |

*Model estimates for Questions 3 and 4 are calculated using the population level model described in section 2.1, 'Model description'.

**Table S11. Comparison of symptomatic COVID-19 cases prevented by Pfizer COVID-19 vaccine versus cases of Pfizer vaccine-associated myocarditis under different intensities of community transmission.** Assuming transmission of delta variant; 5% of unvaccinated, 5% received first dose, 60% received two doses; 30% received three doses; vaccine effectiveness against symptomatic infection as reported in Table S1; age-sex-specific myocarditis incidence as shown in Table S7.

| Age group (years) | Community transmission intensity (probability of infection over 2 months) | Estimated COVID-19 cases over 2 months (per million[a]) | | Estimated COVID-19 cases prevented over 2 months if 5% had 1st dose, 60% had 2 doses, 30% had three doses (per million[a]) | Estimated cases of vaccine-associated myocarditis if 5% had 1st dose, 60% had 2 doses, 30% had 3 doses (per million[a]) | Estimated cases of symptomatic COVID-19 prevented per vaccine-associated myocarditis cases |
|---|---|---|---|---|---|---|
| | | 0% vaccinated | 5% had 1st dose, 60% had 2 doses, 30% had 3 doses | | | |
| **All ages ≥12[b]** | 1% | 9259 | 2378 | 6881 | 28 | 246 |
| | 5% | 46,295 | 11,889 | 34,406 | | 1229 |
| | 10% | 92,590 | 23,777 | 68,813 | | 2458 |
| **12-19** | 1% | 13,768 | 2739 | 11,029 | 91 | 121 |
| | 5% | 68,840 | 13,696 | 55,144 | | 606 |
| | 10% | 137,679 | 27,391 | 110,288 | | 1212 |
| **20-29** | 1% | 13,478 | 3625 | 9853 | 58 | 170 |
| | 5% | 67,390 | 18,125 | 49,265 | | 849 |
| | 10% | 134,780 | 36,249 | 98,531 | | 1699 |
| **30-39** | 1% | 11,536 | 3103 | 8433 | 24 | 351 |
| | 5% | 57,679 | 15,513 | 42,167 | | 1757 |
| | 10% | 115,259 | 31,026 | 84,333 | | 3514 |
| **40-49** | 1% | 9608 | 2522 | 7086 | 16 | 443 |
| | 5% | 48,040 | 12,608 | 35,432 | | 2215 |
| | 10% | 96,081 | 25,216 | 70,864 | | 4429 |
| **50-59** | 1% | 7352 | 1912 | 5441 | 7 | 777 |
| | 5% | 36,762 | 9559 | 27,203 | | 3886 |
| | 10% | 73,523 | 19,118 | 54,405 | | 7772 |
| **60-69** | 1% | 5045 | 1475 | 3570 | 5 | 714 |
| | 5% | 25,224 | 7373 | 17,851 | | 3570 |
| | 10% | 50,448 | 14,746 | 35,702 | | 7140 |
| **≥70** | 1% | 4026 | 1269 | 2757 | 2 | 1379 |
| | 5% | 20,130 | 6347 | 13,783 | | 6892 |
| | 10% | 40,260 | 12,694 | 27,566 | | 13,783 |

[a]Per million population of each age group, or per million of all ages based on population distribution of Australia. [8]
[b]Calculations for all ages based on population distribution of Australia. [8]
